# Exploring the Relationship Between Global Well-being and CO_2_ Emissions

**DOI:** 10.1101/2024.05.23.24307792

**Authors:** Juliet Ware

**Affiliations:** Department of Psychology, University of British Columbia

## Abstract

This study provides an exploratory statistical assessment of the relationship between individual well-being and CO_2_ per capita emissions, and of the mediating factors of this relationship. Building on research which urges sustainable development to integrate subjective well-being, we collected 45 multidisciplinary indices that consider the environmental and human health of every member state of the United Nations (*N* = 196 countries). We are curious to understand whether the relationship between individual happiness and GHG per capita emissions is mediated by the collected indices. Well-being positively and significantly predicts CO_2_ per capita emissions at a global level. Access to adequate water, multidimensional poverty, and gender inequality show evidence of full mediation of this relationship, while seventeen other variables show evidence of partial mediation. These findings provide practical implications for policy recommendations to downscale carbon consumption while ensuring human well-being.

## INTRODUCTION

### The Climate Crisis

Climate change has set our generation into unprecedented action towards mending the harmful impact humans have brought on our natural environment. The primary driver of such impact has been the burning of fossil fuels like coal, oil, and gas, which generate excess greenhouse gas (GHG) emissions into the atmosphere and create a heating effect on the entire planet (United Nations, n.d.). The concentration of GHG in the earth’s atmosphere is directly linked to the average global temperature, a concentration which has been rising steadily since the industrial revolution, and its most abundant source of emissions comes from carbon dioxide (CO_2_) (United Nations, n.d.). Increasing emissions are escalating environmental hazards, threatening clean air, safe drinking water, and nutritious food supply, and costing significant economic resources (World Health Organization, 2024). These consequences are most strongly felt by those who are contributing least to its causes, and who are least able to protect themselves, therefore acting as a threat multiplier (World Health Organization, 2023). We urgently need to downscale our emissions while ensuring the resilience of human health in vulnerable areas of the world. This requires masses of interdisciplinary research and initiatives, to which the field of psychology has much to contribute.

### Well-Being

Integrating well-being into climate studies ensures a holistic, inclusive and practical approach to sustainable development (Iriarte & Musikanski, 2018). Previous research has suggested a global relationship between happiness, development, and carbon emissions across 61 countries: happiness appears to decline when progressing from underdeveloped to medium developed countries, then rises again from medium developed to developed, but with no relationship to carbon emissions (Sulkowski & White, 2016). Similarly, Maddison & Rehdanz (2020) found that next to GDP, a country’s climate is the most important variable explaining cross-country variations in subjective well-being, underlining the magnitude of potential threat to well-being in locations vulnerable to the effects of climate change. Apergis and Majeed (2021) studied greenhouse gas emissions and cross-national happiness for 95 countries between 1995 and 2015, and found emissions to negatively influence happiness. Specifically, the effects of carbon dioxide emissions, risk exposures, and unsafe water quality are detrimental to human and environmental health (Apergis & Majeed, 2021). Thus far, the literature appears to agree that environmental degradation has a negative impact on well-being, while economic development has a positive impact.

Meanwhile, there are a number of studies suggesting that per capita carbon emissions of countries have a positive impact on well-being. Perhaps the most impressive of such findings was the establishment of the carbon intensity of human well-being (CIWB), which calculates the amount of anthropogenic carbon emissions generated per unit of well-being (Jorgenson, 2014). This framework acknowledges the associations between economic development and well-being, and economic well-being and per capita carbon emissions, and therefore establishes a carbon cost to human well-being (Jorgenson, 2014). They find that affluent nations particularly contribute to an increasing CIWB, which is problematic as this perpetuates the disproportionate impacts of climate change (Jorgenson, 2014). Fanning and O’Neill (2019) tested the relationship between carbon-intensive consumptions and human happiness, and found that wellbeing and consumption are linked up to a turning point, after which there is no relationship. Wealthy countries that have crossed this turning point could reduce resource use with little effect on well-being (Fanning & O’Neill, 2019). They proposed a framework of de-growth in affluent areas: if accompanied by redistribution and valuing community and the environment, such societies may downscale consumption while enhancing well-being (Fanning & O’Neill, 2019). However, the aspects of human and ecological development in less affluent areas that must be prioritized in order to achieve low-carbon well-being remains largely unknown.

### Green Space

Urban green space has been identified as an indicator correlated with social factors explaining happiness beyond economic factors (Kwon et al., 2021). However, green space is regulated by climatic predictors and the increasing impacts of climate change (Billie et al., 2023). Also, more developed countries appear to have more urban green space (Billie et al., 2023), and the distribution of green space disproportionately benefits predominantly White and more affluent communities across the world (Wolch et al., 2014). Increasing green space in low-income neighborhoods can lead to gentrification, as it is primarily a strategy for real estate development (Wolch et al., 2014). Green space appears to be a contentious solution, pointing to the need for more inclusive and sustainable approaches in improving well-being.

The current status of the literature points to a direct relationship between human well-being and climate change. The degradation of environments is harmful to human well-being, while actual emissions appear to be beneficial as they are connected to development. Although green space contributes to happiness and to development, it appears to serve the more affluent populations, whose well-being is suggested to have the highest carbon footprint. Furthermore, it remains largely unknown which specific aspects of human and ecological development are most associated with the global relationship between per capita emissions and well-being. This is especially understudied in less developed countries, where the coupling of climate change and well-being is suggested to be significant (Jorgenson, 2014; Fanning & O’Neill, 2019). This underscores the need for a more inclusive and exhaustive framework for analyzing complex global dynamics. The current study aims to identify what mediates the relationship between global climate change and well-being in order to inform climate policy. To answer this question, we tested global well-being and per capita carbon emissions against a wide variety of factors. We collected 45 global variables encompassing environmental, well-being, economic, and socio-political factors and tested their prediction of CO_2_ and well-being. As of yet, no study has been so comprehensive and multidisciplinary in analyzing well-being and climate change. Given the exploratory nature of this project, we did not hypothesize any factors to show prediction or mediation. Instead, we analyze our main findings in the context of recent literature.

## METHODS

### Procedure

We first collected country-level datasets (*N* = 197 countries) from a variety of online databases and sorted them into four categories: environmental, wellbeing, sociopolitical, and economic. The collected datasets are exhaustive: the environmental category includes twenty-eight factors^1^, the wellbeing category includes five factors^2^, the economic category include six factors^3^, and the sociopolitical variable includes five factors^4^. We compiled each variable into one dataset listing the 196 member states of the United Nations. We then computed the relationship between well-being and CO_2_ emissions. To identify mediators, we computed simple linear regressions to test if carbon emissions were predicted by any of our collected variables. With these results, we conducted mediation analyses using the Preacher-Hayes bootstrap method to identify the mediators between countries’ CO_2_ emissions and well-being scores. A detailed description of the source and calculation of each variable, with references, along with the full dataset is available on OSF at the following link: https://osf.io/9swf8/?view_only=16e1a1e1409c4b2f835a5d1076f53470

### Participants

Due to the wide variety of indices collected, the people from which each index is calculated varies greatly. Many indices result from household surveys (e.g. Well-being Index), which reflect the general population in each country. Some indices are calculated by experts in each country (e.g. Electoral Democracy Index), while others are purely geographical (e.g. Global Surface Water Change). Therefore, our collected sample is diverse. Specific demographic information, if applicable, is available in the description of each variable on OSF. Since this research used publicly available data through various online databases, ethics approval was not required. This study did not collect any human participants, rather it used information which had previously been collected from different sources, so participant consent was not required. Researchers did not have access to any identifiable information of individual participants during or after data collection, as data was fully anonymized at the time of collection. This online data was accessed between September and December of 2023, and publication dates of the individual indices are available on OSF.

## RESULTS

### Well-being

We conducted a log transformation on the per capita emissions variable, and found that global well-being is positive, significant, and linear when plotted against log CO_2_ (*B=*.491, *SE*=.053, *t*=9.278, *p*<.001, *R*^2^adj. =.375) (See Figure 1). The well-being score is captured based on answers to life evaluation questions of six factors: economic production, social support, life expectancy, freedom, absence of corruption, and generosity (World Happiness Report, 2022). Due to the lack of emotive components to this index (i.e., the emotional quality of an individual’s everyday experience), we established that this index captures subjective well-being (i.e. the evaluation of overall someone’s life satisfaction). This identified relationship was at the basis of the subsequent mediation analyses in each category.

**Figure 1.**
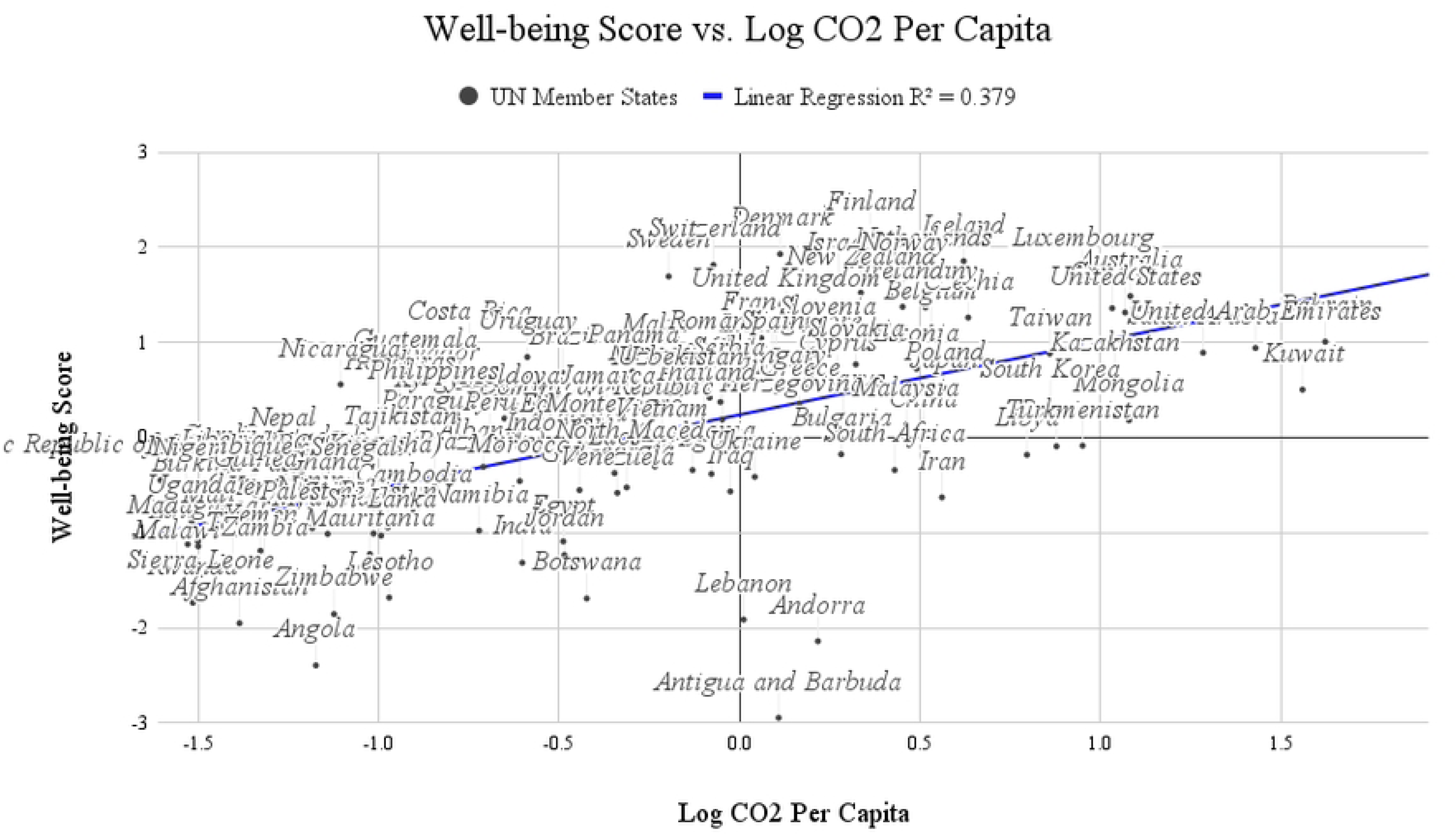
Global Well-being Score plotted against LogCO_2_ per capita emissions. Figure 1 illustrates the distribution of all UN member-states on a coordinate plane, where the upper-right quadrant displays the countries with high carbon emissions and high well-being, and the lower-left quadrant displays the countries with low carbon emissions and low well-being. The upper-left quadrant displays the countries which have achieved high well-being and low carbon emissions. There are not many countries in the lower-right quadrant, which indicates low well-being and high carbon emissions.

#### 1. Environmental Category

We collected GHG emission profiles for each country in 2021, and narrowed the data to include only national CO^2^ emissions measured in million tonnes, and per capita CO^2^ emissions measured in tonnes per capita (Our World in Data, 2021). We used only per capita results in our regression analyses. Table 1.1 lists the results of each factor in the environmental catgeory’s prediction of CO^2^ per capita emissions, ranked in order of the strongest *R*^2^_adj_.

**Table 1.1.** Regression analyses for each index in the environmental category.

| Index | Description | B | SE | t | $R^2_{adj}$ |
| --- | --- | --- | --- | --- | --- |
| Water Quality (access) | Monitors safe water supply by measuring the access to an improved water source that is free from contamination. | .628*** | .047 | 13.319 | .607 |
| EPI: Water Quality (health) | Captures the quality of water in terms of age-standardized disability-adjusted life-years lost per 100,000 persons due to exposure to unsafe drinking water and unsafe sanitation. | .623*** | .040 | 15.622 | .584 |
| Indoor Air Pollution (health) | Number of deaths attributed to indoor air pollution. | -.579*** | .042 | -13.728 | .497 |
| Planetary Pressures | Adjusts the Human Development Index for planetary pressures, which reflects carbon and material footprint. | .421*** | .059 | 7.082 | .243 |
| Environmental Performance Index (EPI) | Measures the green level of a country and determines the level of environmental protection set by its laws, policies, and services, by aggregating 40 different environmental metrics. | .379*** | .054 | 6.988 | .211 |
| EPI: Ecosystem Vitality | Measures how well countries are preserving, protecting, and enhancing ecosystems and the services they provide. Includes six categories (biodiversity & habitat, ecosystem services, fisheries, acid rain, agriculture, and water resources). | .274*** | .058 | 4.763 | .108 |
| EPI: Recycling | Calculates the proportion of recyclable materials (metal, plastic, paper, and glass) recycled in each country. | .256*** | .058 | 4.414 | .094 |

| Index | Description | B | SE | t | $R^2_{adj.}$ |
| --- | --- | --- | --- | --- | --- |
| Renewable Subsidies | Represents government spending toward renewable energy production and consumption. Measured in \$ billion USD. | .190** | .063 | 3.008 | .046 |
| Climate Risk Index (CRI) | How impacted each country is by extreme weather events, by averaging the absolute number of deaths, number of deaths per 100,000 people, absolute losses in PPP, and losses per GDP unit. | .169** | .060 | 2.809 | .037 |
| Non-Renewable Subsidies | Represents government spending toward fossil fuel energy production and consumption. Measured in \$ billion USD. | .166* | .063 | 2.616 | .034 |
| Air Pollution | Monitors how the yearly average PM2.5 concentration ($\mu\text{g}/\text{m}^3$) meets WHO guidelines. | -.154* | .071 | -2.161 | .030 |
| EPI: Policy Objective | Monitors the policy commitments made by the nations in the Glasgow Climate Pact. | .132* | .060 | 2.186 | .021 |
| Ambient Air Pollution (Ozone) | Death rate from air pollution: an estimated annual number of deaths attributed to ambient ozone air pollution per 100,000 people. | -.129* | .059 | -2.190 | .020 |
| Terrestrial Protected Area | Areas of at least 1,000 hectares that are designated by national authorities in order to preserve their ecosystem services and cultural values. Excludes marine areas. | .114 | .059 | 1.934 | .014 |
| Deforestation | Deforestation driven by imported goods minus exported goods. | .093 | .062 | 1.497 | .007 |
| Freshwater Protected Area | The proportion of freshwater Key Biodiversity Areas (KBAs) which are covered by designated protected areas. | .018 | .064 | 0.274 | -.006 |
| Air Pollution (health) | Death rate from air pollution: an estimated annual number of deaths attributed to all causes from air pollution per 100,000 people. | .012 | .059 | 0.195 | -.005 |
| Forest Cover | Measures land with natural or planted stands of trees at least five meters in height, whether productive or not, and excludes tree stands in agricultural production systems. | -.021 | .059 | -0.361 | -.005 |
| Marine Protected Area | Captures the average share of each Marine Key Biodiversity Area that is protected. | .083 | .067 | 1.226 | .004 |
| Surface Water Change (permanent to seasonal water) | Characterises surface water changes from 1984 to 2015 at monthly intervals. Quantifies % of surface water gain and loss in terms of permanent water to seasonal water. | .041 | .060 | 0.693 | -.003 |
| Red List Index | Shows extinction risk for species within the country. Based on data from IUCN Red list of threatened species. | .075 | .059 | 1.269 | .003 |
| Ambient Air Pollution (PM) | Death rate from air pollution: an estimated annual number of deaths attributed to ambient particulate matter air pollution per 100,000 people. | .044 | .060 | 0.733 | -.002 |
| Surface Water Change (seasonal water to permanent water) | Characterises surface water changes from 1984 to 2015 at monthly intervals. Quantifies % of surface water gain and loss in terms of seasonal water to permanent water. | -.053 | .060 | -0.886 | -.001 |
| Surface Water Change (permanent to no water) | Characterises surface water changes from 1984 to 2015 at monthly intervals. Quantifies % of surface water gain and loss in terms of permanent water to not water. | .063 | .060 | 1.058 | .001 |
| Surface Water Change (no water to permanent water) | Characterises surface water changes from 1984 to 2015 at monthly intervals. Quantifies % of surface water gain and loss in terms of no water to permanent water. | -.057 | .060 | -0.956 | -.000 |

*Note: \* $p < 0.05$ ; \*\* $p < 0.01$ ; \*\*\* $p < 0.001$ .*

Water Quality: We ran a linear regression with 5 different metrics included in the EPI (mitigation objective, policy objective, recycling, ecosystem vitality, and water quality), and only the water quality index positively and significantly predicted CO_2_ per capita emissions. This strong predictor might therefore explain the identified relationship between CO_2_ per capita emissions and the EPI. Furthermore, we measured access to water from a different database, and found another positive and significant prediction. It seems an increase in CO_2_ per capita emissions is paired with an increase in access to adequate drinking water.

Air Quality: Although we collected numerous identifiers of air pollution, we found that indoor air pollution most negatively and significantly predicts CO_2_ per capita emissions. We also found that ozone and PM2.5 air pollution significantly and negatively predict CO_2_ per capita emissions. It seems as though various forms of air pollution are negatively coupled with per capita emissions.

We next ran mediation analyses using JASP in order to determine whether these significant predictors mediate the identified relationship between CO_2_ per capita emissions and well-being. Table 1.2 lists the results of the Preacher-Hayes bootstrap mediation method.

**Table 1.2.** Mediation analyses for each significant predictor of the environmental category.

| Mediator | Direct effects | Indirect effects | Total effects |
| --- | --- | --- | --- |
| Water Quality (Health) | B=-.001, $p=.990$, 95% CI (-.232, .211) | B=.714, $p<.001$, 95% CI (.554, .937) | B=.713, $p<.001$, 95% CI (.586, .860) |
| Water Quality (Access) | B=.177, $p=.169$, 95% CI (-.188, .431) | B=.586, $p<.001$, 95% CI (.339, .910) | B=.763, $p<.001$, 95% CI (.621, .908) |
| Indoor Air Pollution | B=.533, $p<.001$, 95% CI (.324, .734) | B=.262, $p=.012$, 95% CI (.090, .424) | B=.794, $p<.001$, 95% CI (.650, .942) |
| EPI | B=.475, $p<.001$, 95% CI (.325, .616) | B=.214, $p<.001$, 95% CI (.146, .345) | B=.716, $p<.001$, 95% CI (.583, .861) |
| EPI: Ecosystem Vitality | B=.596, $p<.001$, 95% CI (.452, .758) | B=.143, $p<.001$, 95% CI (.072, .240) | B=.738, $p<.001$, 95% CI (.601, .870) |
| Planetary Pressures | B=.494, $p<.001$, 95% CI (.258, .657) | B=.259, $p<.001$, 95% CI (.159, .479) | B=.753, $p<.001$, 95% CI (.599, .910) |
| Non-Renewable Subsidies | B=.803, $p<.001$, 95% CI (.656, .961) | B=-.025, $p=.188$, 95% CI (-.069, .005) | B=.778, $p<.001$, 95% CI (.643, .924) |
| EPI: Recycling | B=.665, $p<.001$, 95% CI (.518, .810) | B=.090, $p=.033$, 95% CI (.038, .169) | B=.755, $p<.001$, 95% CI (.608, .899) |
| Renewable Subsidies | B=.839, $p<.001$, 95% CI (.697, .984) | B=-.055, $p=.034$, 95% CI (-.109, -.021) | B=.784, $p<.001$, 95% CI (.655, .934) |
| CRI | B=.786, $p<.001$, 95% CI (.639, .939) | B=-.015, $p=.453$, 95% CI (-.064, .020) | B=.771, $p<.001$, 95% CI (.625, .917) |
| Air Pollution | B=.673, p<.001, 95% CI (.510, .822) | B= .091, p=.032, 95% CI (-.010, .228) | B=.764, p<.001, 95% CI (.626, .912) |
| Policy Objective | B=.702, p<.001, 95% CI (.573, .864) | B= .043, p=.073, 95% CI (.005, .115) | B=.745, p<.001, 95% CI (.613, .903) |
| Ambient Air Pollution (Ozone) | B=.752, p<.001, 95% CI (.633, .909) | B= .016, p=.266, 95% CI (-.007, .061) | B=.768 p<.001, 95% CI (.642, .930) |

**Table 2.1.** Regression analyses for each index in the wellness category.

| Index | Description | B | SE | t | R<sup>2</sup><sub>adj.</sub> |
| --- | --- | --- | --- | --- | --- |
| Life Expectancy | Represents the mortality levels of the population of any country. Measured in years in 2023 by the UN. | .516*** | .047 | 10.967 | .386 |
| Happiness | Calculates scores based on answers to life evaluation questions, over six factors (economic production, social support, life expectancy, freedom, absence of corruption, and generosity) to explain differences between countries. | .491*** | .053 | 9.278 | .375 |
| Education | Creates an index from the expected years of schooling with the mean years of schooling. | .474*** | .049 | 9.731 | .346 |
| Human Rights Index | Represents the extent to which people are free from government torture, political killings, and forced labor; they have property rights; and enjoy the freedoms of movement, religion, expression, and association. | .157** | .062 | 2.528 | .030 |
| Happy Planet Index | Measure of sustainable well-being using limited environmental resources. Key drivers of wellbeing are being active, connecting, taking notice, keeping learning, and giving (none of which have to cost the earth) measured against resource consumption. | .036 | .069 | 0.525 | -.005 |

Note: \* $p < 0.05$ ; $p < 0.01$ ; \*\*\* $p < 0.001$ .

**Table 2.2.**
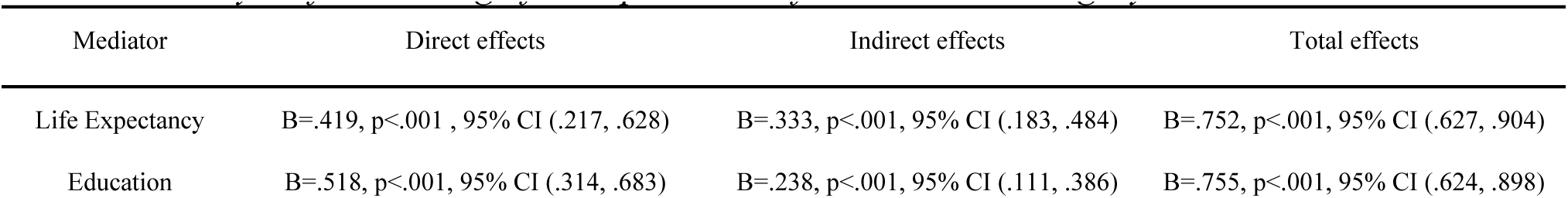
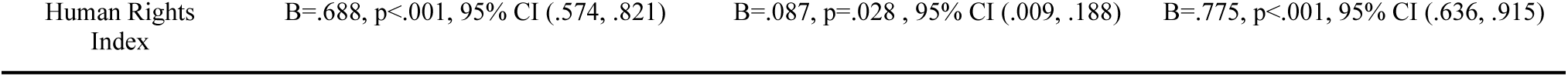
Mediation analyses for each significant predictor of the wellness category.

Both indices of water quality show maximum evidence of a full mediation of the relationship between happiness and GHG per capita emissions, with a non-significant direct effect and significant indirect effect. Indoor Air Pollution, Environmental Performance Index and Ecosystem Vitality each show partial mediation, with a significant direct and indirect effect. The rest of the variables do not show evidence for mediation. Among the 25 collected variables in the environmental category, only water quality and access appear to fully mediate the relationship between carbon impact and well-being.

#### 2. Wellness Category

Every variable in this category except for the Happy Planet Index significantly predicts CO_2_ per capita emissions. We ran Preacher-Hayes bootstrap mediation analyses on each of the significant predictors of GHG per capita emissions to determine any mediators in the wellness category of the relationship between well-being and CO_2_ emissions.

None of the variables shows maximum evidence for full mediation. However, all three show partial mediation, with significant direct and indirect effects. There is not one index of well-being which was found to have a very close association to the carbon impact of well-being, although life expectancy, education, and human rights are partially involved in the relationship.

#### 3. Economic Category

This category includes any variable which reflects the economic health of a country. Table 3.1 lists the simple linear regression results from the economic variables predicting GHG per capita emissions.

**Table 3.1.**
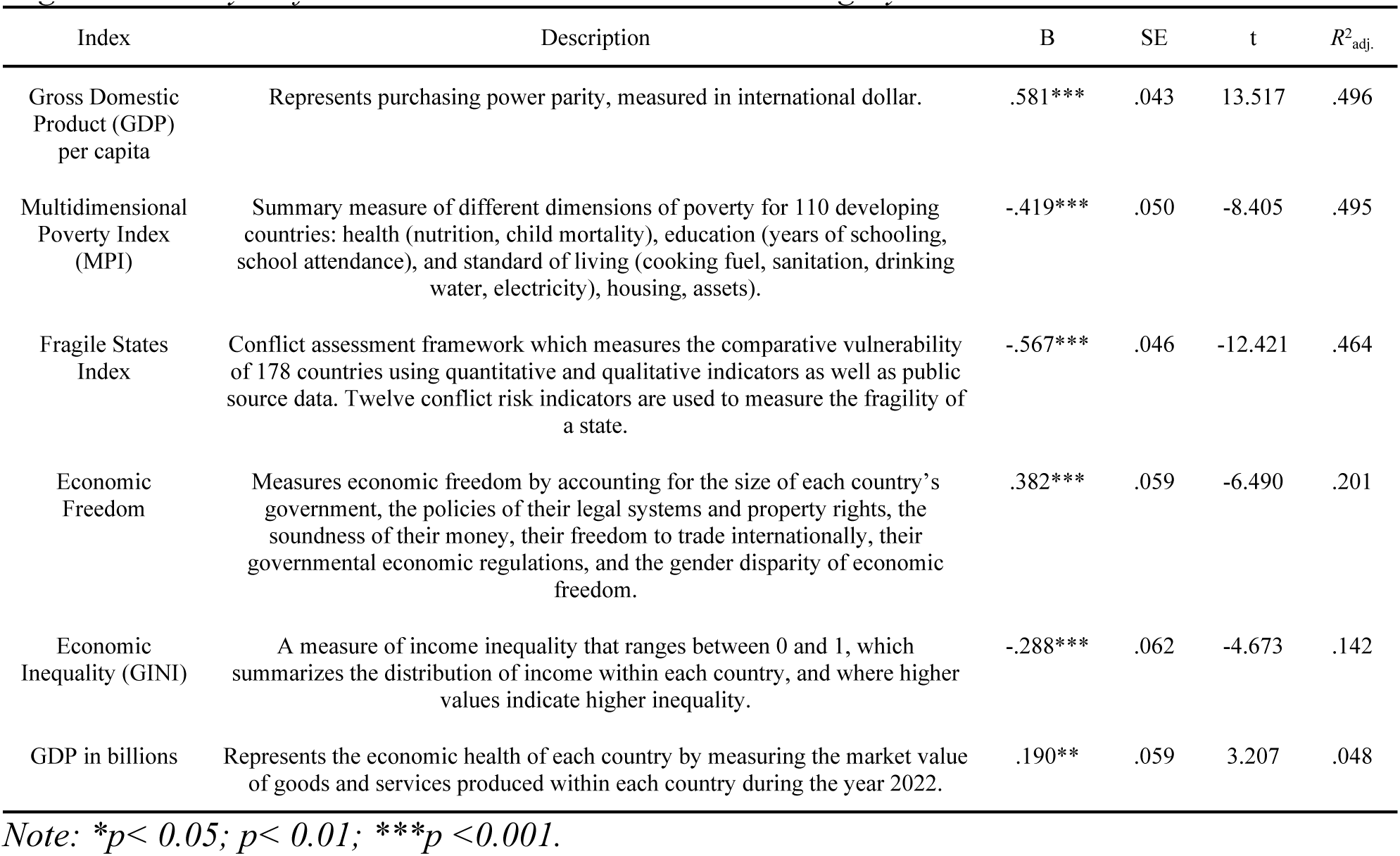
Regression analyses for each variable in the economic category.

| Index | Description | B | SE | t | $R^2_{adj}$ |
| --- | --- | --- | --- | --- | --- |
| Gross Domestic Product (GDP) per capita | Represents purchasing power parity, measured in international dollar. | .581*** | .043 | 13.517 | .496 |
| Multidimensional Poverty Index (MPI) | Summary measure of different dimensions of poverty for 110 developing countries: health (nutrition, child mortality), education (years of schooling, school attendance), and standard of living (cooking fuel, sanitation, drinking water, electricity), housing, assets). | -.419*** | .050 | -8.405 | .495 |
| Fragile States Index | Conflict assessment framework which measures the comparative vulnerability of 178 countries using quantitative and qualitative indicators as well as public source data. Twelve conflict risk indicators are used to measure the fragility of a state. | -.567*** | .046 | -12.421 | .464 |
| Economic Freedom | Measures economic freedom by accounting for the size of each country's government, the policies of their legal systems and property rights, the soundness of their money, their freedom to trade internationally, their governmental economic regulations, and the gender disparity of economic freedom. | .382*** | .059 | -6.490 | .201 |
| Economic Inequality (GINI) | A measure of income inequality that ranges between 0 and 1, which summarizes the distribution of income within each country, and where higher values indicate higher inequality. | -.288*** | .062 | -4.673 | .142 |
| GDP in billions | Represents the economic health of each country by measuring the market value of goods and services produced within each country during the year 2022. | .190** | .059 | 3.207 | .048 |

Note: \* $p < 0.05$ ; $p < 0.01$ ; \*\*\* $p < 0.001$ .

Every collected variable in the economic category significantly predicts CO_2_ per capita emissions. Economic factors are thus quite closely tied to the carbon impact of countries. We examined mediation analyses for each variable; Table 3.2 lists the results.

**Table 3.2:** Mediation analyses for each significant predictor in the economic category.

| Mediator | Direct effects | Indirect effects | Total effects |
| --- | --- | --- | --- |
| MPI | B=.261, p=.234, 95% CI (.235, .720) | B=.512, p=.014, 95% CI (.107, .967) | B=.773, p<.001, 95% CI (.621, .916) |
| Fragile States Index | B=.213, p=.012, 95% CI (.036, .346) | B=.538, p<.001, 95% CI (.405, .701) | B=.750, p<.001, 95% CI (.621, .895) |
| GDP per capita | B=.274, p=.007, 95% CI (.057, .475) | B=.465, p<.001, 95% CI (.320, .678) | B=.740, p<.001, 95% CI (.610, .883) |
| Economic Freedom | B=.505, p<.001, 95% CI (.355, .672) | B=.263, p<.001, 95% CI (.170, .392) | B=.769, p<.001, 95% CI (.636, .919) |
| GINI | B=.659, p<.001, 95% CI (.495, .812) | B=.104, p=.015, 95% CI (.024, .208) | B=.763, p<.001, 95% CI (.632, .911) |
| GDP in billions | B=.767, p<.001, 95% CI (.602, .920) | B=.003, p=.848, 95% CI (-.020, .064) | B=.771, p<.001, 95% CI (.613, .911) |

**Table 4.1.** Regression analyses for each variable in the sociopolitical category.

| Index | Description | B | SE | t | R<sup>2</sup><sub>adj.</sub> |
| --- | --- | --- | --- | --- | --- |
| Gender Inequality Index | Composite metric of gender inequality using three dimensions: reproductive health, empowerment and the labor market. Ranges from 0-1, where a low GII value indicates low inequality between women and men. | -.623*** | .042 | -14.751 | .563 |
| Perceived Electoral Integrity | Comprehensive analysis of how well countries' electoral processes meet international standards of electoral integrity. | .364*** | .055 | 6.649 | .207 |
| LGBT Policy Index | Captures the extent to which people outside traditional sexuality and gender categories have the same rights as straight and cisgender people. Combines 18 individual policies. Higher values indicate more rights. | .252*** | .057 | 4.395 | .090 |
| Electoral Democracy Index | Represents the extent to which political leaders are elected by voting rights in free and fair elections with guaranteed freedoms of association and expression. | .231*** | .061 | 3.783 | .071 |
| 2021 Population | (Our World in Data, 2021). | .029 | .059 | 0.484 | -.004 |

Note: \* $p < 0.05$ ; $p < 0.01$ ; \*\*\* $p < 0.001$ .

**Table 4.2.** Mediation analyses for each significant predictor in the sociopolitical category.

| Mediator | Direct effects | Indirect effects | Total effects |
| --- | --- | --- | --- |
| Gender Inequality | B=.004, p=.965, 95% CI (-.220, .219) | B=.697, p<.001, 95% CI (.513, .918) | B=.701, p<.001, 95% CI (.561, .860) |

| Index |  |  |  |
| --- | --- | --- | --- |
| Perceived Electoral Integrity (PEI) | B=.477, p<.001, 95% CI (.306, .614) | B=.270, p<.001, 95% CI (.176, .407) | B=.747, p<.001, 95% CI (.615, .876) |
| LGBT Policy Index | B=.550, p<.001, 95% CI (.419, .680) | B=.164, p<.001, 95% CI (.085, .288) | B=.713, p<.001, 95% CI (.581, .857) |
| Electoral Democracy Index | B=.613, p<.001, 95% CI (.474, .713) | B=.150, p=.001, 95% CI (.053, .276) | B=.763, p<.001, 95% CI (.626, .896) |

Only the Multidimensional Poverty Index shows maximum evidence for full mediation, with non-significant direct effects and significant indirect effects. Every other variable except GDP in billions shows evidence for partial mediation, with significant direct and indirect effects. Thus, every variable collected in the economic category is involved in the relationship between carbon impact and human well-being, although poverty is a specifically strong association.

#### 4. Socio-Political Category

The final category includes any political or social datasets that we collected which describe different political stances between countries.

The Gender Inequality Index, the Perceived Electoral Integrity, the LGBT Policy Index, and the Electoral Democracy Index significantly predict CO_2_ per capita emissions. The status of equality and democracy clearly influences countries’ carbon pollution.

The Gender Inequality Index shows maximum evidence for mediation, with a non-significant direct effect and a significant indirect effect. Each of the other variables show evidence for partial mediation, with a significant direct and indirect effect. Equality and electoral freedom are evidently involved in the carbon impact and well-being relationship, and gender equality shows the strongest association here.

**Table 5.** Results from all regression and mediation analyses.

| Category | Full Mediations | Partial Mediations | No Mediation | No Prediction |
| --- | --- | --- | --- | --- |
| Environmental | Water Quality and Access | Indoor Air Pollution<br>EPI<br>Ecosystem Vitality<br>Planetary Pressures<br>Non-Renewable Subsidies<br>Recycling<br>Renewable Subsidies | CRI<br>PM2.5 Air Pollution<br>Policy Objective<br>Ozone Air Pollution | Terrestrial Protected Area<br>Deforestation<br>Freshwater Protected Area<br>Air Pollution (all causes)<br>Forest Cover<br>Marine Protected Area<br>Surface Water Change<br>Red List Index<br>Ambient Air Pollution |
| Wellness |  | Life Expectancy<br>Education<br>Human Rights Index |  | Happy Planet Index |
| Economic | Multidimensional Poverty Index | Fragile States Index<br>GDP per capita<br>Economic Freedom<br>GINI | GDP in billions |  |
| Sociopolitical | Gender Inequality Index | Perceived Electoral Integrity<br>LGBT Policy Index<br>Electoral Democracy Index |  | 2021 Population |

## DISCUSSION

Our analyses yielded many significant results. We identified three full mediations, seventeen partial mediations, and eighteen variables which do not mediate or predict the relationship between well-being and CO_2_ emissions. Across all categories, we identified one environmental, one economic, and one sociopolitical variable which show full mediation of the relationship between global well-being and per capita CO_2_ emissions: Water Quality, Multidimensional Poverty Index (MPI), and Gender Inequality. These are three factors which show a close prediction of the direction and geographies in the changing rates of well-being and carbon emissions between countries around the world. These are important and interesting findings: the most recent World Happiness Report (Helliwell et al., 2023) does not mention water quality or access nor gender equality once, and it links poverty to the fragility of the state, which discredits the intricate composition and state of poverty. Thus, gender, water, and poverty are not sufficiently addressed as important drivers of well-being, as is established in our results. The following discussion engages with previous literature to explain our main findings.

### Poverty

The MPI consists of a summary measure of different dimensions of poverty: health (nutrition, child mortality), education (years of schooling, school attendance), and standard of living (cooking fuel, sanitation, drinking water, electricity, housing, assets) (UNDP, 2023). A study by Strotmann and Volkert (2018) investigated which of the MPI indicators have the strongest effects on well-being. They studied 2300 individuals in a rural area of India with the aim of fostering rural development, and identified three aspects of the composite index to predict happiness: education, standard of living, and “missing dimensions” (Strotmann and Volkert, 2018). Participants living in a household in which no member has completed at least 5 years of schooling show a lower probability of being happy, as well as those living in deprivation with respect to ownership of assets and flooring (Strotmann and Volkert, 2018). The “missing dimensions” includes aspects of deprivation that are not captured in the MPI, and they found shame, agency, and empowerment to show significant correlations with well-being (Strotmann and Volkert, 2018). Among the composite index, it appears as though education and empowerment over ownership and agency are strongly connected to well-being in developing areas. These are useful indicators to take into account when considering poverty reduction for the sake of well-being. Moreover, insights from political ecology indicate that poverty specifically affects the livelihoods of women, based on gendered divisions of labor, knowledge, legal rights, and land and natural resource access (Rocheleau et al., 1996 as cited in Ergas and York, 2012). It is important to take into account that these implications of poverty on well-being are disproportionately experienced by women around the world.

Our analyses reveal that a decrease in poverty is coupled with increased well-being and environmental impact. In other words, countries that have lower rates of poverty are more well-off but contribute more to climate change. This is consistent with research establishing that economic development has resulted in a decrease in absolute poverty worldwide, but with a significant increase in global emissions (Malerba, 2020). Researchers have developed a framework for analyzing this trend: the Carbon Intensity of Poverty Reduction (CIPR), a composite indicator that integrates measures for both poverty and environmental outcomes (Malerba, 2020). They found that the CIPR is heterogeneous, explained by economic growth. Up to a certain point, it seems that economic growth reduces poverty without contributing to climate change, but as countries become more developed their carbon consumption and pollution increases (Malerba, 2020). They also identified inequality to reduce the CIPR, which aligns with the body of literature suggesting that changes in inequality play a statistically significant role in decreasing poverty levels (Malerba, 2020). This research confirms our findings that poverty is connected to climate change, and further explains that the pursuit of economic growth in richer countries is not an environmentally sustainable solution to reduce or eradicate poverty. Rather, reducing inequality might be the most synergistic way to increase well-being without furthering climate change. Similarly, Kahneman and Deaton (2010) identified that life satisfaction is strongly correlated with income, but that the positive experience of emotion is only correlated with income up to a point of 75,000$. These findings underscore the importance of reducing suffering from poverty, rather than enhancing well-being in more affluent areas.

Our findings suggest that poverty is tightly connected to climate change and well-being. In line with previous literature, it seems as though this coupling is due to economic growth and efforts to eradicate poverty which unfortunately have a negative impact on the environment. Research on pathways for sustainable models of poverty reduction are needed, though we can identify that reducing inequality, increasing education, and promoting the empowerment and agency of affected individuals in developing areas are low carbon-impact means to reduce poverty and enhance human well-being.

### Water Quality and Access

Our findings establish a close relationship between water quality, carbon emissions, and well-being. The index we gathered measures water quality in terms of age-standardized disability-adjusted life-years lost per 100,000 persons due to exposure to unsafe drinking water and unsafe sanitation (Yale, 2022). According to our analyses, the countries with the most highly contaminated water are also the most unhappy and the least contributing to climate change. Furthermore, we found that a separate metric measuring access to safely managed drinking water also fully mediates this relationship. This assesses access to an improved water source (piped water, public taps, tube wells, dug wells, protected springs, rainwater collection) located on premises, available when needed and free from contamination. Our analyses reveal that improved access to clean water is found in countries with higher carbon emissions and higher well-being. Relative to all other variables collected in the environmental category, the quality of water and its accessibility are the most highly important when considering climate change and well-being.

UNICEF (2023) establishes that 1 in 4 people around the world lack safely managed drinking water, although between 2000-2022, 2.1 billion people gained access and the number of people lacking access decreased. However, ⅔ of those who gained access lived in urban areas. The need for adequate access for clean water is therefore crucial in rural areas. To foster sustainable development, low-carbon solutions to improve water conditions in such areas are needed to increase well-being without contributing to climate change.

Guardiola, González-Gómez, and Grajales (2013) tested the influence of water on well-being in Yucatan, Mexico. This area experiences low quality water access and imperfections in their water management services (e.g. water cuts). They found that in Yucatan, the water domain of life has a significant and positive effect on well-being, while money, quality of house, leisure, and community do not (Guardiola et al., 2013). Among the determinants of the water domain, they identified perceived water quality to have a positive effect, and purchase of bottled water to have a negative effect, while having a tap or well and water cuts have no effect (Guardiola et al., 2013). Nadeem et al. (2020) ran a similar experiment in the rural farmlands of Faisalabad, Pakistan, which faces serious water quality and access issues due to industrial pollution, water systems, and agriculture. In Faisalabad, the likelihood of water scarcity driven by climate change alone is greater than 90% (Nadeem et al., 2020). These researchers identified that the source and quality of drinking water, access to irrigation water, % of crop water, and water expenses all have an influence on the well-being of rural farmers (Nadeem et al., 2020). Another element of the relationship between water and well-being was identified by Marcantonio (2018), who studied the Choma district of Zambia and found that the perception of waterborne illness is tightly coupled with well-being. 33% of the observed sample reported illness from drinking water, and this was identified to have a negative effect on happiness (Marcantonio, 2018). Additionally, women globally experience more waterborne illnesses and other health risks because they are most responsible for water procurement (WHO, 2023). The lack of access to clean water and hygiene therefore disproportionately affects women and girls around the world as they are primarily responsible for domestic chores and care work (WHO, 2023). Essentially, researchers have successfully identified that water is a very important component to well-being, particularly affecting vulnerable rural areas and women.

Access to adequate water is a significant problem that is likely to escalate due to worsening floods, rising sea levels, shrinking ice fields, wildfires, and droughts as a result of climate change (UN, nd.). Water solutions must factor in well-being, as our analyses have established this to be a close relationship. However, our findings also identify a close relationship between improved water conditions and increased CO_2_ emissions. There is a call for additional research here to reduce the carbon intensity of water development, while prioritizing human well-being. Evidence from rural areas in Mexico, Pakistan and Zambia recommend improving well-being by reducing contamination in wells and taps, revitalizing groundwater for agricultural purposes, and minimizing water expenses – with a specific priority towards female health equity.

### Gender Inequality

Considering the disproportionate burden women and girls face in instances of both poverty and inadequate water conditions, the finding that gender equality alone is a full mediator merits much attention. There is an impressive amount of literature which considers gender equality to be greatly involved in both economic development and carbon emissions. First, to explain our findings: the Gender Inequality Index is a composite metric using three dimensions: reproductive health, empowerment, and the labor market. It reflects gender-based disadvantages and the differences in female versus male achievements (UNDP, 2022). We found gender inequality to reduce where both per capita CO_2_ emissions and well-being increase. Countries with more inequality tend to be less pollutive and less happy, but there is an apparent carbon cost to increasing equality.

This finding can first be understood in the context of a study which found that gender equality increases happiness across developing and emerging countries (Ndoya et al., 2024). This research suggests that gender equality makes our societies richer and more secure, providing transformative effects (Ndoya et al., 2024). Women’s full and active participation in society improves the quality of the workforce and leads to public policies that affect society’s well-being (Ndoya et al., 2024). Qian (2017) identified that among the indicators of the Gender Inequality Index, maternal mortality, education, and sexual empowerment most significantly predict increased happiness among 169 nations. Labor market participation, however, has a partial contribution towards happiness, but the ratio of female to male labor force participation has a negative relationship with happiness (Qian, 2017). It may be that this heightened economic vitality from labor force participation is also contributing to countries’ per capita CO_2_ emissions.

Ergas et al. (2021) explored the effect of gender equality on the carbon intensity of well-being (CIWB), and found that labor force participation tends to increase emissions and reduce life expectancy. Women are often hired in industry work which is dangerous and highly pollutive (i.e. sweatshops), and this might contribute to increasing environmental impact. However, they also identified that women’s educational attainment and proportion of legislative positions significantly reduce emissions, countering this effect (Ergas et al., 2021). The contribution of gender equality on carbon impact is thus two-fold: labor participation increases CIWB, but legislative and educational empowerment reduce CIWB. Moreover, McGee et al. (2020) studied the moderating effect of gender inequality on economic development and carbon impact across 140 nations. They found that gender inequality strengthens the relationship between economic growth from CO_2_ emissions across 140 nations, and that this is especially strong in less developed countries (McGee et al., 2020). Higher inequality is especially damaging to the environment in less developed areas. However, women’s political empowerment and educational attainment weakens this relationship (McGee et al., 2020; Ergas et al., 2021). This aligns with the well-documented finding that female executive participation on company boards around the world has a significant negative impact on carbon emissions and carbon disclosure (Zhang et al., 2024; Issa and In’airat, 2024; Abd Majid and Jaaffar, 2023; Kim, 2022).

There are multiple dynamics at play which influence the interaction between gender equality, environmental impact, well-being, and human development. This area of research is new and ongoing, and must continue to parse out the specific mechanisms involved in order to provide adequate policy recommendations. Our findings establish that gender equality mediates the relationship between well-being and carbon impact, and this might be explained by multiple reasonings. To summarize, female health, education, and sexual empowerment greatly contribute to well-being. However, female labor force participation tends to reduce well-being and increase emissions, unless attenuated by an increase in legislative and executive positions held by women. This may be due to the tighter coupling of economic development and carbon pollution when there is high inequality in less developed countries. To strengthen this reasoning, income was identified as a partial mediator in the current study, suggesting it has an influence in this relationship. The decoupling of gender inequality and environmental impact is especially important in developing countries. Our results and analysis can provide the following recommendations: developing countries would benefit from policies that focus on the educational attainment of women and on implementing labor protection that center the safety and health of women joining the workforce; all countries would benefit from an increase in female leadership and empowerment in executive roles.

## CONCLUSION

After examining the 45 variables of the global dataset, access to adequate water, multidimensional poverty, and gender equality have the most significant influence on the relationship between global CO_2_ emissions per capita and well-being. Multiple other multidisciplinary variables show evidence of partial mediation, although we do not interpret these findings in detail. The implications of the current study connect to a global desire for healthier and happier people and environment. Changes in our understanding of global well-being are required in order to reduce its impact. Currently, well-being seems to be largely grounded in economic development: efforts to decouple this relationship are required to enhance human well-being without furthering the climate crisis. Our research suggests directions and areas of work which might be central to this decoupling. Our findings provide recommendations to reduce the carbon impact of well-being. First, female leadership, health, and education empowerment can provide low-carbon means of achieving greater gender equality and reducing poverty. Second, reducing contamination of groundwater and wells in rural and developing areas have low-impact and support areas most affected by poor water quality. Third, improving costs of clean water can sustainably improve its accessibility and human well-being. However, such recommendations arise from the current state of literature on the intersecting fields of climate change and global inequalities, which is new and unfinished. Much more research is needed to provide effective adaptation and mitigation plans. We also recommend analyzing the partial mediators identified in this data analysis as they are significant predictors of well-being and carbon impact and can inform further policy implications.

Our study was limited by the different online databases where we sourced our information. Most sources did not include every country within the United Nations, so we worked through many missing data points. Additionally, the sources collected do not come from the same year, although they are all recent. We recommend future research includes geographic factors which might have an influence on the relationship between well-being and CO_2_ emissions. For example, countries could be coded for Global North/South, Western/non-Western, or coastal/regional. We also recommend analyzing similar variables across time. Lastly, we recommend using statistical clustering to identify the specific countries most vulnerable to poverty, inadequate water conditions, and inequality. This would allow specific targeting and case studies for improving well-being without furthering climate change.

## Data Availability

All data files are available from the OSF database (https://osf.io/9swf8/?view_only=16e1a1e1409c4b2f835a5d1076f53470).

## Acknowledgements

I would like to acknowledge my research supervisor Dr. Jiaying Zhao. I could not have taken on this work without her continual support and guidance.

## Footnotes

1 Environmental variables include: national CO2 emissions, CO2 emissions per capita, Climate Risk Index, Environmental Performance Index, Mitigation Performance, Policy Objective Performance, Recycling Performance, Ecosystem Vitality, Water Quality Index, Access to Safely Managed Drinking Water, Air Pollution, Deaths Caused by Air Pollution – including indoor, particulate matter, and ozone pollution –, Planetary Pressures, Non-Renewable Energy Subsidies, Renewable Energy Subsidies, Red List Index, Measures of Surface Water Change, Terrestrial Protected Area, Forest Cover, Marine Protected Area, Freshwater Protected Area, and Net Deforestation in Trade.

2 Well-being variables include: Happiness score, Happy Planet Index, Life Expectancy, Human Rights Index, and Mean Education in years.

3 Economic variables include: GDP (billions USD), GDP per capita (international dollars), Economic Freedom Summary Index, GINI coefficient, Multidimensional Poverty Index, and Fragile States Index.

4 Sociopolitical variables include: Electoral Democracy Index, Gender Inequality Index, LGBT+ Policy Index, PEI Index, and population.

## Notes

### Competing Interest Statement

The authors have declared no competing interest.

### Funding Statement

The author(s) received no specific funding for this work.

### Author Declarations

This study did not require ethics approval.

